# Trends in Thoracic Impedance and Arrhythmia Burden Among Patients with Implanted Cardiac Defibrillators During the COVID-19 Pandemic

**DOI:** 10.1101/2021.02.27.21252559

**Authors:** Yuan Lu, Karthik Murugiah, Paul W Jones, César Caraballo, Shiwani Mahajan, Daisy S Massey, Rezwan Ahmed, Eric M Bader, Harlan M Krumholz

## Abstract

Hospitalizations for acute cardiac conditions have markedly declined during the coronavirus disease 2019 (COVID-19) pandemic, yet the cause of this decline is not clear. Using remote monitoring data of 4,029 patients with implantable cardiac defibrillators (ICDs) living in New York City and Minneapolis/Saint Paul, we assessed changes in markers of cardiac status among these patients and compared thoracic impedance and arrhythmia burden in 2019 and 2020 from January through August. We found no change in several key disease decompensation markers among patients with implanted ICD devices during the first phase of COVID-19 pandemic, suggesting that the decrease in cardiovascular hospitalizations in this period is not reflective of a true population-level improvement in cardiovascular health.

## INTRODUCTION

During the coronavirus disease 2019 (COVID-19) pandemic, hospitalizations for acute cardiac conditions including heart failure have markedly declined.^1, 2^ This may be because patients are avoiding seeking care during the pandemic or because cardiovascular events are fewer due to changes in behaviors or exposures. Implantable cardiac defibrillators (ICDs) implanted in patients with heart failure provide useful information on markers of cardiac status as measured by thoracic impedance, arrhythmia burden, and device discharges.^3^ Using remote monitoring data of patients with ICDs living in New York City (NYC) and Minneapolis/Saint Paul (MSP), we sought to assess changes in markers of cardiac status among these patients and compared thoracic impedance and arrhythmia burden in 2019 and 2020 from January through August.

## METHODS

A de-identified dataset of individuals from NYC and MSP aged ≥18 years with ICD devices transmitting between January 1^st^, 2019 and August 22^nd^, 2020 was derived from the Boston Scientific Corporation’s LATITUDE™ database. Measured variables included daily thoracic impedance, nighttime heart rate, respiration rate, heart rate variability, time in atrial fibrillation (AF), percentages of patients with AF time greater than 1 hour, and HeartLogic index for heart failure decompensation. For this analysis, we used remote monitoring data exclusively from scheduled data uploads to eliminate potential bias in event-based data uploads.

We assessed the trend in daily thoracic impedance, arrhythmia burden, and device discharges between January 1^st^, 2020 and August 22^nd^, 2020. We used data during the same period from 2019 as a baseline to account for seasonal variability and then calculated the change in these variables during and after COVID-19 restrictions in 2020. Year to year comparisons for the same time period were performed using a repeated measures ANOVA. We considered 2-sided P-values <0.05 as statistically significant. Analyses were performed using R version 4.0.2 (CRAN institute). This study was approved by the Institutional Review Board at Yale University.

## RESULTS

A total of 4,029 patients were included in the analysis, of whom 2,792 were from NYC (mean age 67.9 [SD, 13.5] years; 31.9% women) and 1,237 were from MSP (mean age 67.3 [SD, 13.3] years; 28.9% women). There was little to no statistical or clinically meaningful change in thoracic impedance and arrhythmia burden in patients with ICD devices during the COVID-19 shut-down period (**Figure**). The average daily nighttime heart rate, heart rate variability, and HeartLogic index for heart failure decompensation decreased slightly (relative change less than 5%) from the date that the emergency quarantine order was issued (03/07/20 in NYC and 03/13/20 in MSP) to the end of August. Conversely, average daily thoracic impedance, AF, and percentages of patients with daily AF time greater than 1 hour increased slightly in the same time period. However, none of these changes were statistically significant except for the decrease of heart rate variability in NYC and the increase of AF burden in MSP. Similar trends were seen in both NYC and MSP and were consistent across age and sex subgroups.

## DISCUSSION

There is little to no change in most markers for disease decompensation among heart failure patients with implanted ICD devices in NYC and MSP during the COVID-19 pandemic, and this is consistent for all age and sex subgroups. The lack of changes in cardiac status among a high-risk group suggests that the decline in inpatient cardiovascular services was not a result of marked improvement in cardiovascular health.

There may be several explanations for the lack of change. First, the effects of behavioral changes on disease decompensation are not immediate. As such, a brief period of behavioral changes such as reduced physical activity due to lockdown may not result in immediate changes in markers of cardiac status. Second, the effects of behavioral changes may be offset by changed exposures. For example, the loss of physical activity may be compensated for by shifts in dietary patterns and reduced exposure to ambient stressors.

These findings have implications regarding the causes of the decrease in cardiovascular hospitalizations during the shutdown. Media reports expressed concerns that people were avoiding care.^4, 5^ An alternative hypothesis is that cardiovascular health improved with better air quality and fewer risk behaviors. Our study fails to provide evidence that cardiovascular health improved (or worsened) among people with cardiovascular disease. These findings instead necessitate health care attention and highlight the importance of ensuring high-risk patients have access to critical clinical care during the current COVID-19 pandemic.

Limitations of our study include inability to ascertain clinical outcomes, the lack of information on other factors that may affect disease decompensation markers, and the short follow-up observation period.

In conclusion, we found no change in several key disease decompensation markers among patients with implanted ICD devices during the first phase of COVID-19 pandemic, suggesting that the decrease in cardiovascular hospitalizations in this period is not reflective of a true population-level improvement in cardiovascular health. We will need to remain vigilant to prevent a second pandemic of neglected cardiovascular disease in the coming months.

## Data Availability

Aggregate data of the results are available upon request to authors.

## FIGURE LEGENDS

**Figure.**
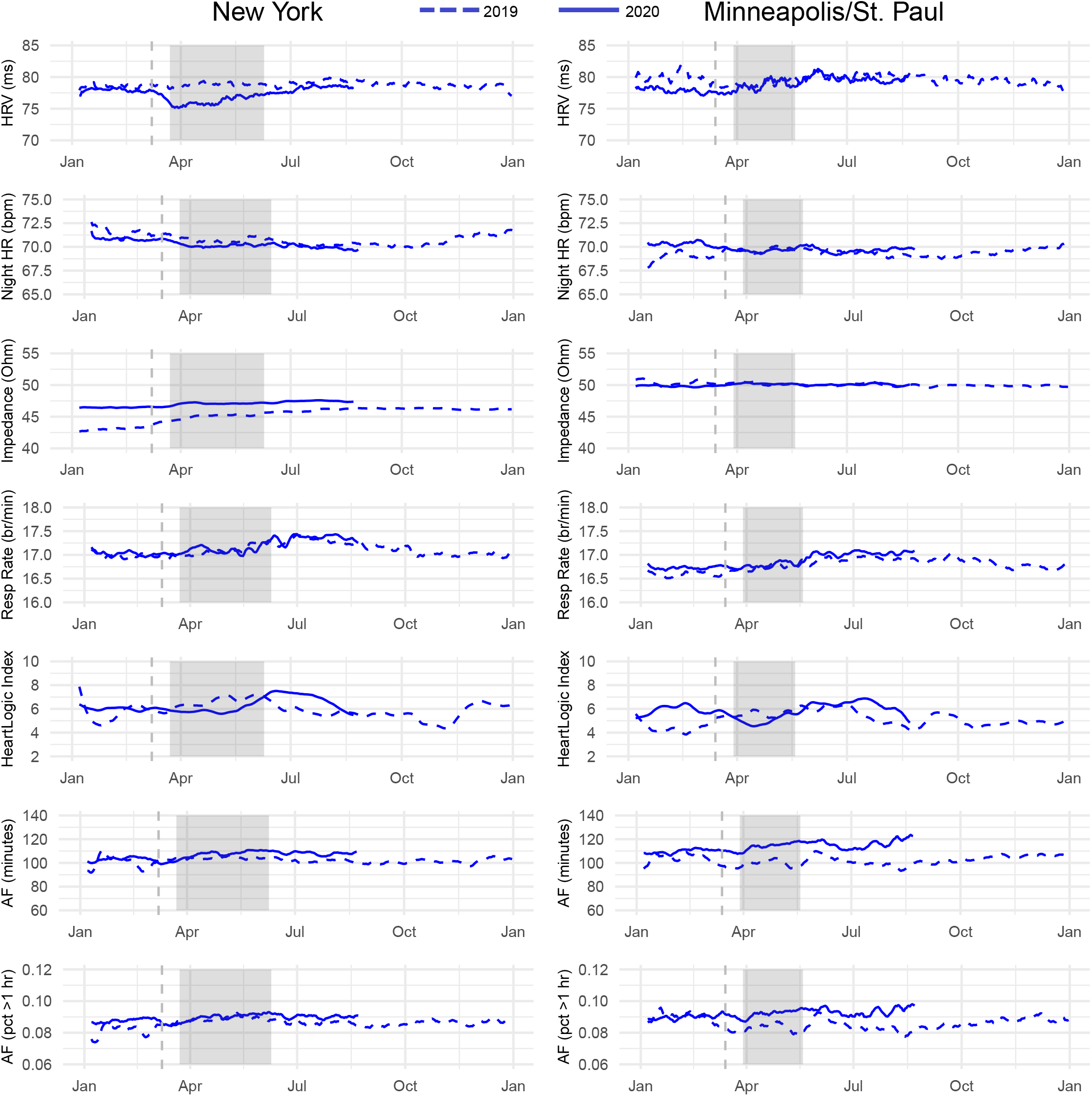
Trends in thoracic impedance and arrhythmia burden among patients with implanted cardiac defibrillators in New York City and Minneapolis/Saint Paul in 2019 and 2020. The left side is New York City, and the right side is Minneapolis/Saint Paul. The dashed line is 2019 and the solid line is 2020. The vertical line is the date of emergency order and the shaded area is the stay-at-home order for each city. Abbreviations: HR, heart rate; HRV, heart rate variability; Resp rate, respiration rate; AF, atrial fibrillation.

## REFERENCES

1. Bhatt AS, Moscone A, McElrath EE, Varshney AS, Claggett BL, Bhatt DL, Januzzi JL, Butler J, Adler DS, Solomon SD and Vaduganathan M. Fewer hospitalizations for acute cardiovascular conditions during the COVID-19 pandemic. Journal of the American College of Cardiology. 2020;76:280–288.

2. De Filippo O, D’Ascenzo F, Angelini F, Bocchino PP, Conrotto F, Saglietto A, Secco GG, Campo G, Gallone G, Verardi R, Gaido L, Iannaccone M, Galvani M, Ugo F, Barbero U, Infantino V, Olivotti L, Mennuni M, Gili S, Infusino F, Vercellino M, Zucchetti O, Casella G, Giammaria M, Boccuzzi G, Tolomeo P, Doronzo B, Senatore G, Grosso Marra W, Rognoni A, Trabattoni D, Franchin L, Borin A, Bruno F, Galluzzo A, Gambino A, Nicolino A, Truffa Giachet A, Sardella G, Fedele F, Monticone S, Montefusco A, Omede P, Pennone M, Patti G, Mancone M and De Ferrari GM. Reduced rate of hospital admissions for ACS during Covid-19 outbreak in northern Italy. The New England journal of medicine. 2020;383:88–89.

3. Parthiban N, Esterman A, Mahajan R, Twomey DJ, Pathak RK, Lau DH, Roberts-Thomson KC, Young GD, Sanders P and Ganesan AN. Remote monitoring of implantable cardioverter-defibrillators: A systematic review and meta-analysis of clinical outcomes. Journal of the American College of Cardiology. 2015;65:2591–2600.

4. Cigna study finds reduced rates of acute non-elective hospitalizations during the COVID-19 pandemichttps://www.cigna.com/about-us/newsroom/studies-and-reports/deferring-care-during-covid-19. Accessed on October 20, 2020.

5. Krumholz H. Where have all the heart attacks gone? https://www.nytimes.com/2020/04/06/well/live/coronavirus-doctors-hospitals-emergency-care-heart-attack-stroke.html. Accessed on October 20, 2020.

